# UDCA May Promote COVID-19 Recovery: A Cohort Study with AI-Aided Analysis

**DOI:** 10.1101/2023.05.02.23289410

**Authors:** Yang Yu, Guo Yu, Lu-Yao Han, Jian Li, Zhi-Long Zhang, Tian-Shuo Liu, Ming-Feng Li, De-Chuan Zhan, Shao-Qiu Tang, Zhi-Hua Zhou, Guang-Ji Wang

**Affiliations:** National Key Laboratory for Novel Software Technology, Nanjing University, China; School of Basic Medicine and Clinical Pharmacy, China Pharmaceutical University, China; Hospital of Nanjing University, Nanjing University, China; State Key Laboratory of Natural Medicines, Key Laboratory of Drug Metabolism, China Pharmaceutical University, China; Polixir.ai

**Author notes:** Co-first authors (Yang Yu, Guo Yu, Lu-Yao Han, Jian Li, Zhi-Long Zhang, Tian-Shuo Liu).

## Abstract

To investigate the impact of ursodeoxycholic acid (UDCA) treatment on the clinical outcome of mild and moderate COVID-19 cases, a retrospective analysis was conducted to evaluate the efficacy of UDCA on patients diagnosed with COVID-19 during the peak of the Omicron outbreak in China. This study presents promising results, demonstrating that UDCA significantly reduced the time to Body Temperature Recovery after admission and a higher daily dose seems to be associated with a better outcome without observed safety concerns. We also introduced VirtualBody, a physiologically plausible artificial neural network model, to generate an accurate depiction of the drug concentration-time curve individually, which represented the absorption, distribution, metabolism, and excretion of UDCA in each patient. It exhibits exceptional performance in modeling the complex PK-PD profile of UDCA, characterized by its endogenous and enterohepatic cycling properties, and further validates the effectiveness of UDCA as a treatment option from the drug exposure-response perspective. Our work highlights the potential of UDCA as a novel treatment option for periodic outbreaks of COVID-19 and introduces a new paradigm for PK-PD analysis in retrospective studies to provide evidence for optimal dosing strategies.

---

The COVID-19 pandemic has caused an enormous global burden on public health, affecting civil societies, and hindering economic development ^1^. In China, the number of COVID-19 infections skyrocketed in November 2022, following the government’s active optimization and refinement of its COVID-19 response. The most prevalent strains were BA.5.2 (70.8%) and BF.7 (23.4%) ^2,3^. Therefore, given the emergence of new variants and the persistent risk of periodic outbreaks during the era of Omicron, the need for accessible, oral therapeutic options to treat COVID-19 remains urgent, especially for individuals who have already been vaccinated ^4–6^. Currently, only two oral antivirals, ritonavir-boosted nirmatrelvir and molnupiravir, have been approved for the treatment of mild or moderate COVID-19 under Emergency Use Authorization ^7–10^. However, their availability is predominantly limited to individuals with affluent financial resources, and their efficacy against the Omicron (B.1.1.529) variant in vaccinated patients is not well established.

Recent studies have explored the potential of UDCA, a classic FXR inhibitor, as a treatment option for COVID19. In a study published in Nature, Brevini et al. reported that FXR plays a crucial role in regulating ACE2 expression, and this regulation can be effectively inhibited in vitro using organoids and *ex situ* on a pair of human lungs by UDCA ^11^. Furthermore, they observed a significant association between UDCA treatment and improved clinical outcomes after SARS-CoV-2 infection in a retrospective cohort of liver transplant recipients. However, the current evidence supporting the use of UDCA in the treatment of COVID-19 is based on clinical trials with small sample sizes, which cannot be considered conclusive ^12^.

Between December 10, 2022, and December 30, 2022, during the peak of the Omicron outbreak in China, a cohort of non-severe COVID-19 patients was admitted to Nanjing University Hospital, with some receiving treatment with UDCA and a control group receiving standard care. Subsequently, a retrospective cohort investigation was conducted using deidentified data to assess the effectiveness of UDCA treatment in non-severe COVID-19 patients. Additionally, a subgroup analysis was performed to comprehensively evaluate the impact of various daily doses of the treatment, while accounting for demographic features, concomitant drug use, and clinical baseline on admission.

While this retrospective study provides some insights into the potential effectiveness of UDCA as a treatment, the unexpected emergence of the COVID-19 pandemic poses significant challenges for well-designed, prospective clinical trials. Many COVID-19 drugs result in suboptimal therapeutic outcomes due to poor pharmacokinetic properties or high inter-individual variability ^13,14^. Of note, the enterohepatic cycle profile of UDCA, coupled with its endogenous nature, complicates its pharmacokinetic properties such that using dosage as a substitution of exposure may not be appropriate ^15^. Therefore, additional information acquisition and dose-exposure-response correlation are crucial for the optimization of the UDCA therapeutic strategies in a large population as a novel treatment option for COVID-19.

To address this issue, we present a groundbreaking approach to this challenge with the first, to our knowledge, learning-based high-precision digital human PK-PD model, VirtualBody. This model allows for the accurate derivation of personalized longitudinal PK data and distribution across multiple organ systems in the human body. In addition, the PK parameter area under the curve (AUC) calculated from the simulated pharmacokinetic data not only enhances our understanding of the therapeutic efficacy of UDCA against COVID-19 but also facilitates accurate estimation of its effective concentration. These results provide valuable insights for tailoring UDCA treatment to specific patients, showing a promising avenue for individual dosing regimens optimization.

## Retrospective data analysis

### Real-world data and cohort characteristics

In this retrospective study, 120 patients were diagnosed with COVID-19 between December 10, 2022, and December 30, 2022, of which 5 patients were not hospitalized despite having a fever for four or more days. Following the exclusion criteria, a total of 115 patients were retained for the univariate recovery analysis with adequate individual clinical data and documented diagnosis of COVID-19, 65 of whom had a claim for ursodeoxycholic acid (UDCA), while 50 did not receive any UDCA treatment. Among the UDCA-treated group, 29 (44.6%) and 36 (55.4%) patients received daily doses of 150mg and 300mg or more, respectively. However, due to missing demographic data from 8 patients, the final multivariate adjusted analysis was performed on a subset of 107 patients (Extended Data Figure 1). The baseline characteristics of the different cohorts and subgroups were summarized in Extended Data Table 1-2.

It is worth noting that the patients included in this study were young individuals without any comorbidities, and all had received at least two doses of the COVID-19 vaccine. Although UDCA has been proposed as a potential treatment for COVID-19^11^, the available data is limited to retrospective analyses on patients with cholestasis, progressive liver disease, cirrhosis, or liver transplantation, which are confounded by numerous effects related to the complexity of the target FXR. Specifically, gene mutations and differences in bile acid profiles among patients with different types of cholestasis can affect FXR expression, making it more vulnerable ^16–18^. Additionally, studies have shown that the bile acid composition after UDCA treatment differs between healthy individuals and those with primary biliary cholangitis (PBC) ^19,20^. Therefore, it is crucial to evaluate the effectiveness of UDCA in healthy individuals while controlling for comorbidities and eliminating confounding factors.

### UDCA may promote the body temperature recovery of COVID-19

Among the 115 patients who were included in the univariate analysis (65 treated with UDCA and 50 without UDCA), 61 patients (93.8%) who received UDCA and 38 patients (76.0%) who did not receive UDCA had their body temperature returned to 37.3^°^C or lower before discharge. Moreover, the median recovery time for the UDCA group was 4 days (95% CI, 4-4), compared to 5 days (95% CI, 4-5) for the no-UDCA group. The difference in the distribution of recovery speed between the two groups was significant (*P* = 0.030), as shown in Figure 1a.

**Figure 1:**
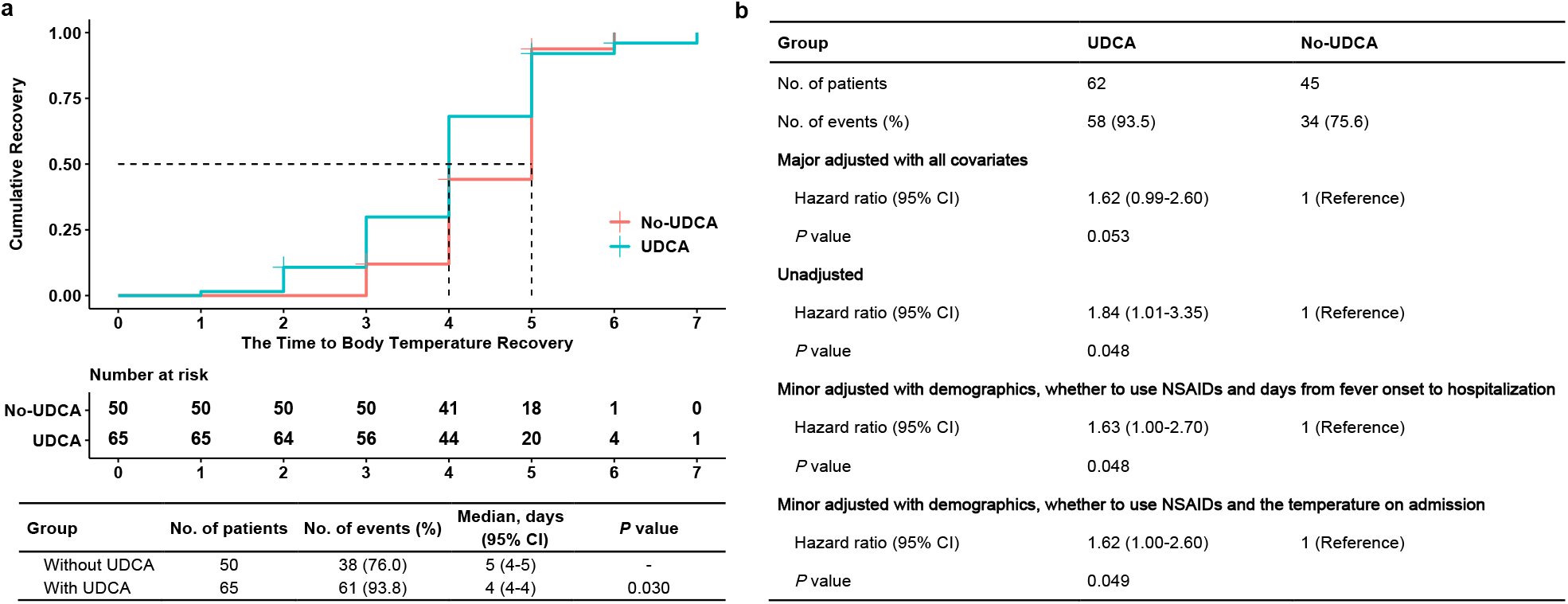
Univariate and multivariate analyses of UDCA and no-UDCA groups. **a**, Univariate analysis of UDCA and no-UDCA groups. **b**, Multivariate analyses of UDCA and no-UDCA groups with major adjusted, unadjusted and different minor adjusted.

**Figure 2:**
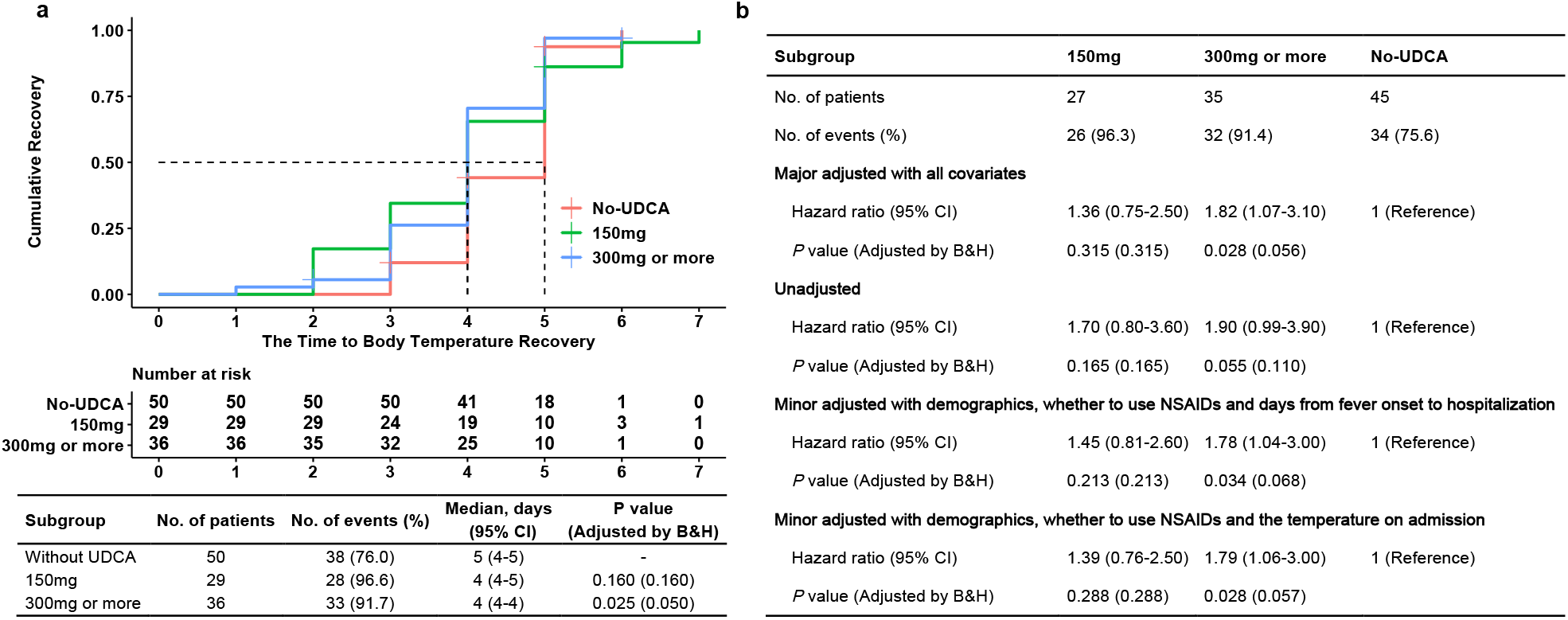
Univariate and multivariate analyses of subgroups. **a**, Univariate analysis of subgroups. **b**, Multivariate analyses of subgroups with major adjusted, unadjusted and different minor adjusted.

After adjusting for covariates, including age, sex, BMI, concomitant NSAID use, days from fever onset to hospitalization, and temperature on admission, the Cox proportional hazards models showed that UDCA treatment was associated with a 62% higher benefit on the speed of Body Temperature Recovery (HR 1.62, 95% CI 0.99-2.60) compared to the no-UDCA group (Figure 1b). In the unadjusted analysis, the hazard ratio of the UDCA group referring to the no-UDCA group was 1.87 (95% CI, 1.06-3.32). The minor adjusted results of the Cox proportional hazards regression model were consistent with the unadjusted analysis, even after different degrees of adjustments for various covariates, indicating the potential utility of UDCA in improving recovery time with a significant level of confidence. Furthermore, to examine the effect of missing covariates on the results, multivariate analyses with imputation were performed and shown in (Extended Data Table 3).

### High dose performed better on primary endpoint

To provide further evidence for the efficacy of UDCA in treating COVID-19, we conducted a subgroup analysis based on dosage. Patients who received a daily dose of 300mg or more of UDCA showed a significant improvement in recovery time compared to the no-UDCA group, regardless of multiplicity adjustment. In contrast, patients who received a daily dose of 150mg did not show a significant difference in recovery time (Figure 1a).

The multivariate analyses were further conducted using Cox proportional models in two subgroups. Results revealed that the major adjusted hazard ratio for the 300mg or more subgroup was significantly associated with a shorter time to Body Temperature Recovery ([HR], 1.82 [95% CI, 1.07-3.10]) compared to the no-UDCA group (Figure 1b), while the 150mg subgroup did not demonstrate such an association without multiplicity adjusting. Additionally, minor adjustment methods yielded similar results to the primary analysis with major adjustments. Moreover, the estimated hazard ratios of both subgroups in the unadjusted analysis did not show significant differences from the baseline no-UDCA group, with multiplicity adjusting. Multivariate analyses with imputation were shown in (Extended Data Table 4), indicating that the high-dose subgroup had more significant results when the sample size was sufficient.

## AI generated data analysis

Establishing a correlation between UDCA dosage/exposure and efficacy in COVID-19 treatment is difficult to achieve through retrospective data alone. Artificial intelligence techniques can help infer unobserved data. We developed a high-precision digital model of the human body called VirtualBody, which uses physiologically structured neural networks to learn real-world data. We use the generated data from VirtualBody to enhance our analysis.

### Overview of VirtualBody

VirutalBody comprises three integral components: the Human feAtures conditioned Diffusion (HAD) Model, the Skeleton Model, and the Skin Model. These three components underwent pre-training and fine-tuning using their distinct datasets, and their pipelines are shown in Supplementary Materials^1^. Upon the completion of the training pipeline, VirtualBody had the capability to generate individualized PK data and inferred the dissemination of UDCA across multiple organ systems, even with extremely limited clinical data in the general population. Firstly, our approach leveraged high-dimensional human body characteristics that were automatically extracted from HAD Model. These characteristics were inputted into the Skeleton Model, which subsequently rolled out the venous drug concentration and drug concentrations in other organs under a specific dosage protocol autoregressively. Following this step, the Skin Model ultimately utilized the circulation data generated by the Skeleton Model to match the transition distribution of PK data, thus enabling the simulation of various clinical results.

Technically, VirtualBody models the transition of PK as a Markov Decision Process (MDP), defined by a tuple *M* = (*S, A, M* ^∗^, *r, γ, ρ*_0_) ^21^, where *S* is the state space, *A* is the action space, and *M* ^∗^ is the optimal (i.e., real-world) transition model. As for our MDP, it mainly consisted of three key elements:

1. States incorporated detailed information about the drug, including its chemical properties and patientspecific features derived from the HAD Model. Time information is also included to ensure the Markov property is satisfied. In addition to the time-independent features, the dynamic drug concentration transition data is an essential dimension for the states, as our model is optimized to generate drug concentration-time curve.
2. Actions at a given time step were two-dimensional, with the first dimension being the medicine dosage and the second dimension representing patients’ diets.
3. State transitions defined the probability distribution over the next states, i.e. the PK data at the next time point, given a state and an action.

The learning objective of VirtualBody was to predict the subsequent state for each time step within the trajectory in this MDP, based on the corresponding state-action pair at that particular step.

Overall, VirtualBody enabled the handling of missing data during inference, the accommodation of various dosing strategies, and the prediction of venous and organ concentrations at any given time step. It is skilled in adapting to diverse dosage protocols and variations among individuals during inference, thus providing information to change the dosage regimen for the better.

### VirtualBody generates accurate PK data for UDCA

The performance of VirtualBody was compared with the traditional top-down approach PopPK and other supervised learning methods, using longitudinal time-concentration data collected from a bioequivalence trial. The trial involved 120 individuals in a randomized, open-label, two periods, two sequences, crossover, single dose, fasting, and fasted design. The data was collected over a 72-hour period at various time points. The evaluation of PK-PD analysis was performed based on fundamental evaluation indices including Goodness of Fit (GOF), Area Under the Curve (AUC), and Maximum Concentration (C_max_). It was demonstrated that VirtualBody surpassed other baselines in the analysis. The evaluation results are presented in Table.1.

**Table 1:** UDCA evaluation metric for VirtualBody and baselines

|  | Method | Evaluation Metric |  |  |  |
| --- | --- | --- | --- | --- | --- |
|  |  | R Square | MAE | MAPE | Correlation |
| GOF | MLP | - 1.943 ± 2.521 | 1.295 ± 0.374 | — | 0.608 ± 0.098 |
| | LSTM | - 0.354 ± 0.474 | 1.811 ± 0.419 | — | 0.393 ± 0.088 |
| | Transformer | - 0.315 ± 0.399 | 1.117 ± 0.201 | — | 0.619 ± 0.046 |
| | PopPK | - 0.228 ± 0.353 | 1.125 ± 0.053 | — | 0.525 ± 0.110 |
|  | VirtualBody | **0.123 ± 0.166** | **0.820 ± 0.043** | — | **0.627 ± 0.055** |
| AUC<sub>0-72h</sub> | MLP | — | 63.032 ± 48.457 | 2.306 ± 1.287 | — |
| | LSTM | — | 59.754 ± 28.060 | 2.903 ± 1.575 | — |
| | Transformer | — | 27.605 ± 6.840 | 1.238 ± 0.293 | — |
| | PopPK | — | 18.319 ± 2.800 | 0.927 ± 0.248 | — |
|  | VirtualBody | — | **10.368 ± 2.709** | **0.426 ± 0.109** | — |
| $C_{\max}$ | MLP | — | 3.954 ± 0.500 | 0.785 ± 0.292 | — |
| | LSTM | — | 2.882 ± 0.588 | 0.555 ± 0.075 | — |
| | Transformer | — | 2.866 ± 0.921 | 0.499 ± 0.121 | — |
| | PopPK | — | 2.698 ± 0.859 | **0.456 ± 0.044** | — |
| | VirtualBody | — | **2.619 ± 0.268** | 0.469 ± 0.074 | — |

Goodness of Fit (GOF) is a statistical measure that point-wisely evaluates the accuracy of a model in predicting observed drug concentration data. Based on the evaluation results, it is apparent that VirtualBody gained superiority over all other baselines on all metrics, achieving exceptional performance in modeling drug behavior in vivo. Specifically, R-square is a widely used indicator of how well a model fits the data, with a value close to one indicating a good fit. UDCA, being an endogenous bile acid with many uncertain PK properties, generally exhibited a negative baseline R-square. This can be attributed to factors such as accumulation in the bile acid pool, a typical hepatointestinal circulation profile, and susceptibility to the effects of diet and gallbladder contraction. However, a positive R-square (0.123) was achieved by VirtualBody in the prediction of UDCA PK data, making it the only model to our knowledge with this ability, and demonstrating its accuracy in individual variation prediction. Moreover, the outstanding performance of VirtualBody was further evidenced by its low Mean Absolute Error (MAE) and high correlation with the observed data, indicating its impressive ability not only to predict drug concentrations more accurately at each timestep but also to successfully capture the underlying trend of the real data.

In regards to one of the most crucial PK parameters, AUC_0-72h_, VirtualBody outperformed other models with an MAE of only 60% and a MAPE of 50% or even smaller than other models. AUC is a crucial quantitative parameter for describing the total drug exposure over time in vivo, taking into account various factors in the ADME process. This parameter aided in establishing a more precise exposure-efficacy model, usually with less variability in comparison to the dose-efficacy relationship. With respect to C_max_, it is noteworthy that although the MAPE of VirtualBody was slightly larger than that of PopPK, VirtualBody still achieved the lowest MAE. Remarkably, VirtualBody had the minimum standard deviation in almost all indicators, indicating that VirtualBody was able to maintain high robustness with accurate prediction.

These phenomena indicated that VirtualBody not only delivered more precise predictions of blood drug concentration but also offered better stability in its predictions. As for the conventional methodologies for PK/PD analysis, PopPK suffered from time-consuming human adjustments and its embedded nonlinear mixed-effects model had limited expressive capacity. Supervised learning methods, on the other hand, were likely to be data-hungry if we aimed to achieve strong generalization. Additionally, it can be observed that the most accurate inter-subject variation was captured by VirtualBody, as exhibited by its lowest uncertainty.

### PK-PD analysis with VirtualBody

VirtualBody served as a crucial intermediary between pharmacokinetics and pharmacodynamics by precisely forecasting individual pharmacokinetic profiles, guaranteeing a comprehensive understanding of implicit knowledge in clinical data, and facilitating the discovery of novel insights. Logistic regression analysis was conducted to evaluate the impact of UDCA dose, predicted AUC and AAIC as continuous variables on the likelihood of body temperature recovery. These analyses were based on a longitudinal expansion dataset from original clinical data combined with predicted results, as shown in the schematic diagram of Figure 3a. The results showed that cumulative dose, AUC, and all of AAIC had statistically significant effects on the event of body temperature recovery, with odds ratios greater than 0, after the adjustment for the days from fever onset. However, the odds ratio of AUC was higher than that of dose, indicating a stronger correlation between AUC and febrile events (Figure 3b). Moreover, by increasing the minimum effective concentration of AAIC, there was a corresponding increase in its odds ratio which reached its peak at 1.5*μ*g/mL. This observation highlighted the importance of excluding low-dose AUC and provided practical evidence for UDCA dosage recommendations in clinical administration. The histogram vividly showed that the AUC of patients with body temperature recovery was higher than that of patients without body temperature recovery on the same day after fever, whether in the complete cohort or in UDCA treated group (Figure 3c).

**Figure 3:**
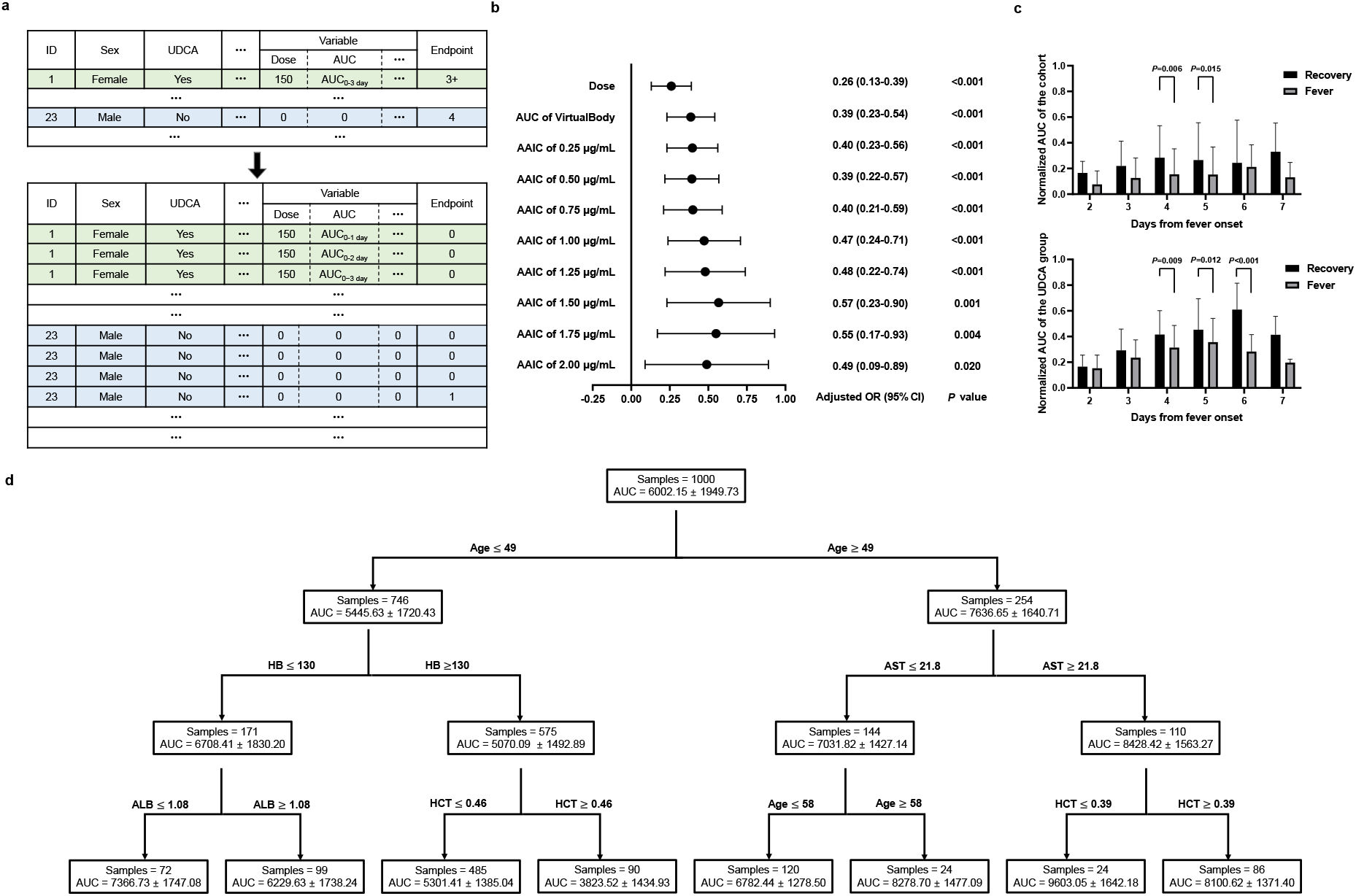
Data expansion and further analysis. **a**, Depiction for longitudinal expansion for each individual. **b**, Logistic regression in longitudinal expansion data. **c**, Histogram for exposure-efficacy relationship in two groups, the AUC bar of corresponding days was shown as Mean*±*SD. **d**, Population separation according to AUC value by decision tree model from a virtual population of 1000 individuals

To illustrate the model’s expansion and identify drug suitability for specific populations, a decision tree was employed to evaluate the impact of various features. Firstly, the impact of age on UDCA exposure was found to be the most significant factor according to the root node in our decision tree. The model detected a substantial difference in average AUC between the two nodes under the age branches. Given UDCA’s high solubility and low permeability, it was more readily absorbed in the elderly due to the slowing of gastrointestinal peristalsis. This finding validated the model’s ability to learn the effects of age on drug absorption and confirmed the pharmacokinetic characteristics of UDCA. Therefore, appropriate dose reduction was recommended for elderly patients receiving UDCA treatment. Population differences were reflected in certain liver function indicators, such as AST and albumin levels. As liver function declined, the AUC of UDCA increased due to its reliance on liver metabolism and transport. This highlighted the sensitivity of UDCA exposure to changes in liver function, with elevated AST and decreased albumin levels resulting in higher AUC values (Figure 3d).

## Discussion

In this retrospective real-world cohort study of 115 hospitalized COVID-19 patients, we have demonstrated that Ursodeoxycholic Acid (UDCA) treatment is significantly associated with a shorter time to Body Temperature Recovery after admission, with intensified effectiveness at a relatively high daily dosage. Our study is one of the first to evaluate retrospectively the real-world effectiveness of UDCA in hospitalized patients with COVID-19, conducted in fully vaccinated patients during the era of the Omicron variant. Our findings demonstrate that UDCA significantly shortened the duration of fever caused by SARS-CoV-2 infection in Chinese patients with non-severe COVID-19, without any observed adverse effects. These promising results warrant further investigation of UDCA as a potential treatment for COVID-19, particularly in healthy individuals without comorbidities. This approach may help mitigate the impact of diseased conditions on targeting the Farnesoid X receptor (FXR) regulation, thereby offering new avenues for effective management of COVID-19.

Although the primary endpoint of our study, the time to Body Temperature Recovery, may not be entirely consistent with other studies, as fever is one of the most common symptoms in patients with COVID-19 infection ^22^, it serves as the main criterion for hospital admission and discharge in the healthcare facilities where our investigation was carried out ^23^.

Exclusion criteria are critical for determining the quality and credibility of research results. In our study, we excluded patients who were not hospitalized after fever for four or more days for two reasons: firstly, the self-limiting characteristics are not uncommon in most non-severe COVID-19 courses, and secondly, the pharmacological mechanism of UDCA inclined to prevent SARS-Cov-2 infection has been clarified. We have demonstrated that the potential of UDCA to modulate Angiotensin Converting Enzyme 2 (ACE2) provides efficacy as both primary and secondary prophylaxis against COVID-19, as it impedes cellular uptake of SARS-CoV-2 through a reduction in binding to the membrane-bound form of ACE2 during the early phase of infection ^24^. Likewise, the efficacy of human recombinant soluble ACE, which has been engineered to specifically bind with SARS-CoV-2 as a substitution of endogenous ACE2, has been proved to significantly reduce viral load ^25^.

While the overall outcome of our study is positive and clinically significant, its weaknesses must be considered. Due to the short observation period, the small sample size, the relatively restricted patient population characteristics, and the lack of a prospective design, the scope and applicability of this investigation are limited. These factors hindered us from carrying out adequate research and undertaking pharmacokinetic-pharmacodynamic (PK-PD) analysis. In the absence of PK/PD profiles, we were unable to establish rational dosages and optimize therapy.

Driven by the data insufficiency, we present the first, to our knowledge, learning-based high-precision digital human PK/PD model called VirtualBody. VirtualBody allows for the estimation of personalized drug distribution throughout the entire human body and PK/PD data with only a small amount of clinical drug trial data from the general population. We demonstrate the effectiveness of our method in successfully modeling the complicated PK/PD data of UDCA. Meanwhile, the skin model utilizes generated circulation data to predict the drug concentration in plasma. Our novel digital model structure represents a significant advancement in the field of clinical trial simulation, with potential applications in personalized medicine and drug development. It offers an efficient and cost-effective auxiliary tool to traditional clinical drug experiments, showing a promising avenue for recognizing the weak points in trial designs and building general human disease models.

In conclusion, our study is the first clinical study conducted in the real world retrospectively evaluating the therapeutic effect after the mechanism of UDCA against COVID-19 was determined. Our study pioneered the use of UDCA in treating COVID-19 infections during the peak of the pandemic in China. As the outbreak of COVID-19 is time-sensitive, our research is invaluable in the real world. This study revealed promising results, demonstrating that the administration of UDCA positively impacted therapeutic outcomes in individuals without comorbidities. Specifically, the UDCA treatment significantly reduced the time required for Body Temperature Recovery following Omicron strain infection, without observed safety concerns. Moreover, using our novel learning-based PK/PD model, VirtualBody, we were able to establish an accurate exposure-efficacy relationship, which complements previous findings based on the dose-efficacy relationship that can be influenced by individual differences in metabolism and drug-drug interactions. In addition, our model provides ample evidence for determining the effective in vivo concentration of UDCA, optimizing the drug dose and dosing regimen, and enhancing the drug’s efficacy and safety. The findings of this study present UDCA as a novel treatment option for periodic outbreaks of COVID-19, paving the way for further large-scale, clinical controlled, and prospective research to explore its full potential.

## Method

### Study design and population

We accessed and reviewed the de-identified data through the Electronic Data Capture system of Nanjing University Hospital in December 2022. The Nanjing University Hospital institutional review boards approved this study as minimal risk and waived informed consent requirements. We excluded patients who experienced fever due to COVID-19 for four or more days ^26^.

### Exposure and endpoints variables

All patients had no previous history of UDCA medication before hospitalization. Patients in the UDCA group were initiated on UDCA treatment on the day of admission, while patients in the no UDCA group did not receive UDCA treatment throughout their hospitalization. We organized patient data of the cohort encompassing demographics (sex, age, BMI), the clinical baseline on admission (days from fever onset to hospitalization and temperature on admission), concomitant drug use (non-steroidal anti-inflammatory drugs, other symptomatic drugs recommended by Chinese National Diagnosis and Treatment of Novel Coronavirus Pneumonia (Version 9) for alleviating symptoms other than fever, such as sore throat, cough, shortness of breath, vomit, and diarrhea), and clinical body temperature recovery. Patients in the UDCA group were assigned to subgroups based on their daily dose, namely 150 mg and 300 mg or more, as only two individuals received 450 mg besides 150 mg and 300 mg.

Our primary endpoint was the time to Body Temperature Recovery, which was defined as the duration it took for body temperature to return to 37.3^°^C or lower after admission and censored on the day of discharge without recovery. For patients with recurrent fever, we recorded the last time of Body Temperature Recovery. The final body temperature for analysis was the measured axillary temperature + 0.5^°^C.

### Preliminary statistical analysis

Baseline characteristics of patients were presented as the median and interquartile range (IQR), unless noted. The differences between UDCA and no UDCA groups in categorical and continuous variables were statistically examined using chi-square tests and Wilcox tests, respectively. All statistical comparisons were two-sided, with significance assumed at *α* = 0.05. *·* Univariate Kaplan-Meier was used to show recovery curves between the two cohorts and subgroups categorized by the daily dose of UDCA, using the primary endpoint as the evaluation measure without adjusting for confounding factors. The median recovery time of the two cohorts and subgroups was estimated with the Brookmeyer-Crowley method, with a 95 % confidence interval. The Log-rank test was applied to compare recovery curves with and without UDCA cohorts. Furthermore, the same method was adapted to compare two subgroups within the UDCA cohort based on their daily dose with the no-UDCA cohort.

To assess the impact of UDCA versus no-UDCA on the time to Body Temperature Recovery, the primary analysis was conducted using Cox proportional hazard model with all covariates adjusted. The covariates available for adjustment include demographical covariates (sex, age, and BMI), concomitant drug use (whether to use NSAIDs), and clinical baseline on admission (days from fever onset to hospitalization and temperature on admission). To avoid bias caused by the selection of covariates, we conducted a sensitivity analysis not adjusting for all covariates, minor adjusting for demographical features, concomitant drug use, and one of the clinical baselines on admission (days from fever onset to hospitalization or temperature on admission). The continuous covariates were normalized to the 0-1 range. The same method was applied to the evaluation between the two dose subgroups and the no-UDCA cohort.

To handle missing covariates, we employed the multiple imputations by chained equations method with ten imputed datasets and ten iterations. The missing covariates were allowed a maximal fraction of missing information (FMI)/m *≤* 10% (Supplementary Table S2). For each covariate, a random forest model was utilized. This method was adopted under the assumption that the missing observations for covariates were missing at random. The validity of the imputations was assessed by comparing the distribution of recorded and imputed values for all measurements through visual inspection. To further evaluate the robustness of our results, we performed a sensitivity analysis on both samples with complete covariates and those with imputed data. The imputation was performed using the mice package. All statistical analyses were performed using R (version 4.2.1). The *P* values of subgroup analyses were presented both before and after Benjamini & Hochberg adjustment.

### The VirtualBody workflow

The model training pipeline was a three-stage framework that comprised feature alignment, VirtualBody pretraining, and VirtualBody finetuning. Firstly, in the feature alignment stage, noticing that there existed a significant gap between the features from PBPK data and the clinical data, we employed a state-of-the-art generative model to align the PBPK features and authentic features, and details were provided in the

### Supplementary Information^2^

Secondly, in the pre-training stage, we separately trained each organ neural network component with pre-collected data, which was encoded into the neural network model by adopting a data-driven approach. We then integrated all the components into the systemic circulation, based on prior physiological mechanisms to form the Skeleton Model. Finally, the fine-tuning stage was to bias the model towards the experimental outputs by utilizing the authentic data and still retaining the pre-trained information in the meanwhile.

### VirtualBody training and validation

There is no denying that the pharmacokinetic process is complex, highly varied among individuals, and influenced by multiple features. During the pre-training state, our goal was for VirtualBody to capture all of the critical prior knowledge.

The authentic UDCA data used for fine-tuning consisted of longitudinal time-concentration data collected from the bioequivalence trial initiated by Azpharm group limited in Guangdong, China (Registration Number: CTR20192232, http://www.chinadrugtrials.org.cn/), and the results of this trial has been reviewed by The Center for Drug Evaluation, National Medical Products Administration, China. The half of 120 subjects received single 500mg ursodeoxycholic acid tablets orally on fasted state, and the other half were administrated the same regimen on the fed state.

A 5-fold cross-validation was conducted on the authentic UDCA dataset to assess the prediction accuracy of the VirtualBody and other baselines. Our evaluation primarily focused on the Goodness of Fit, Area Under the Curve, and Maximum Concentration, to exhibit the comparison of several models from two levels, the fitting of points, and PK parameters.

### Goodness of Fit (GOF)

It was assumed that the total number of drug-time curves was *N*, with each curve consisting of *H* sampling points. The observed concentration and predicted concentration of the *j*-th sampling point of the *i*-th curve were represented by 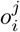 and 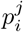. Additionally, 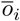 and 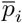 were defined to represent the mean value of the observed concentration and predicted concentration of the *i*-th curve, respectively. The evaluation indicators of GOF were the R square, mean absolute error, and correlation, represented by Eqs. (1), (2), and (3), respectively.

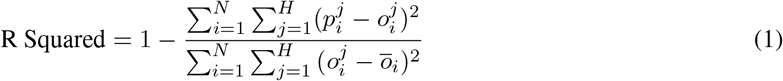

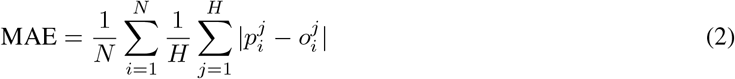

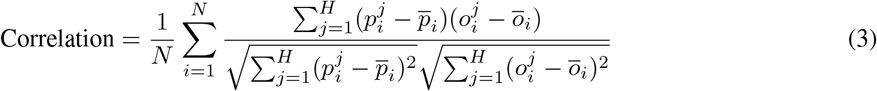

### Area Under the Curve (AUC_0-72h_)

Under similar assumptions in GOF, we defined *ao*_*i*_ and *ap*_*i*_ to represent the observed AUC_0-72h_ and predicted AUC_0-72h_ of the *i*-th curve respectively. The evaluation indicators of AUC_0-72h_ included mean absolute error Eq.(4) and mean absolute percentage error Eq.(5).

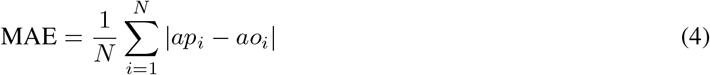

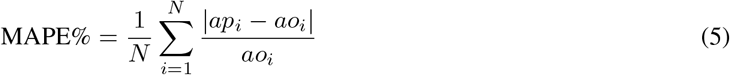

### Maximum of Concentration (C_max_)

Under similar assumptions in GOF, we defined *co*_*i*_ and *cp*_*i*_ to represent the observed maximum concentration and predicted maximum concentration of the *i*-th curve respectively. The evaluation indicators of C_max_) included mean absolute error Eq.(6) and mean absolute percentage error Eq.(7).

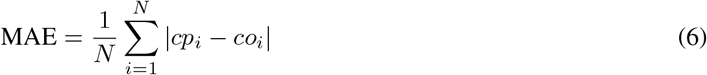

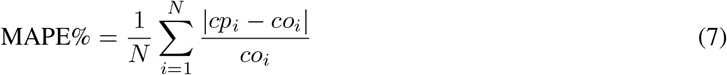

### PK-PD analysis with VirtualBody

Using VirtualBody, we predicted the concentration of UDCA for each sample and calculated the Area Under the Curve (AUC) and the Area Above the Inhibitory Concentration (AAIC) at eight different concentrations ranging from 0.25*μ*g/mL to 2*μ*g/mL. The resulting data were then integrated into the original real-world dataset. In PK-PD analysis, since the baseline of hospitalization in the real-world dataset was misaligned, the time-series data for each individual was divided into multiple samples based on their total duration of fever from the onset, according to the number of days with a fever. Furthermore, each sample was labeled with clinical outcomes as the clinical endpoint to conduct PD analysis, where only the outcome of the non-censored last sample corresponding to temperature recovery was recorded as 1. All other samples and the censored last sample were labeled as 0, indicating that the temperature had not recovered. Logistic regression was performed on the above dataset to investigate the impact of cumulative drug dose or predicted AUC or predicted AAIC on the clinical outcome of temperature recovery. Fever duration, the use of non-steroidal anti-inflammatory drugs, and demographic characteristics of the patients were included as covariates for adjustment.

To gain deeper insights into the capabilities of VirtualBody, we generated a virtual population of 1,000 individuals with a wide distribution of ages ranging from 18 to 60 years old and heights ranging from 1.6m to 1.8m. The subjects were administered a fixed daily dosage of 500mg for 5 days, and VirtualBody was used to predict and calculate the AUC for each individual at the end of the fifth day. Finally, we incorporated a variety of relevant features, such as height, weight, age, sex, and various laboratory test values as input for a decision tree algorithm to predict the AUC for each individual.

## Related work

Pharmacokinetic/Pharmacodynamic (PK/PD) modeling is fundamental for understanding how body organisms interact with drugs through the process of absorption, distribution, metabolism, and finally excretion of the active substance, which is the cornerstone of clinical application ^27,28^ and offer great opportunities to better understand the in vivo behavior of medications in clinically challenging situations ^29^. Now, computational modeling approaches for pharmacokinetics are in two paths that focus on either expert knowledge or experimental data. Focusing on expert knowledge, physiologically based pharmacokinetic (PBPK) modeling and simulation employs differential equations to describe the kinetics in each organ. Population pharmacokinetics (PopPK) is an alternative path that focuses on experimental data. Population pharmacokinetics analysis is a statistical method to explain the variability in drug concentration among individuals ^30^.

Although recent advances in deep learning have shown promising results in various aspects of drug discovery and design, including AlphaFold for protein structure prediction ^31^, DeepDTA macromolecular target identification ^32^ and MiCaM for molecular generation ^33^, the application of deep learning in PK/PD modeling has been limited. The network in neural-PK/PD ^34^ is insufficient to model the PK/PD and not specifically tailored and supervised learning is not well-suited for this task. However, recent successes in applying Deep Reinforcement Learning to challenging decision-making problems in various domains ^35–38^ have shown great potential for modeling drug concentration transitions. A well-trained environment model can improve the sample efficiency, identify optimal policies, and incorporate domain knowledge to explain the policy behavior in the action-predictive decision-making training process ^39,40^. Therefore, DRL with environment model learning provides a data-driven avenue for personalized PK modeling and dosage regimen optimization.

## Data Availability

All data produced in the present study are available upon reasonable request to the authors.

---

**Extended Data Figure 1.**
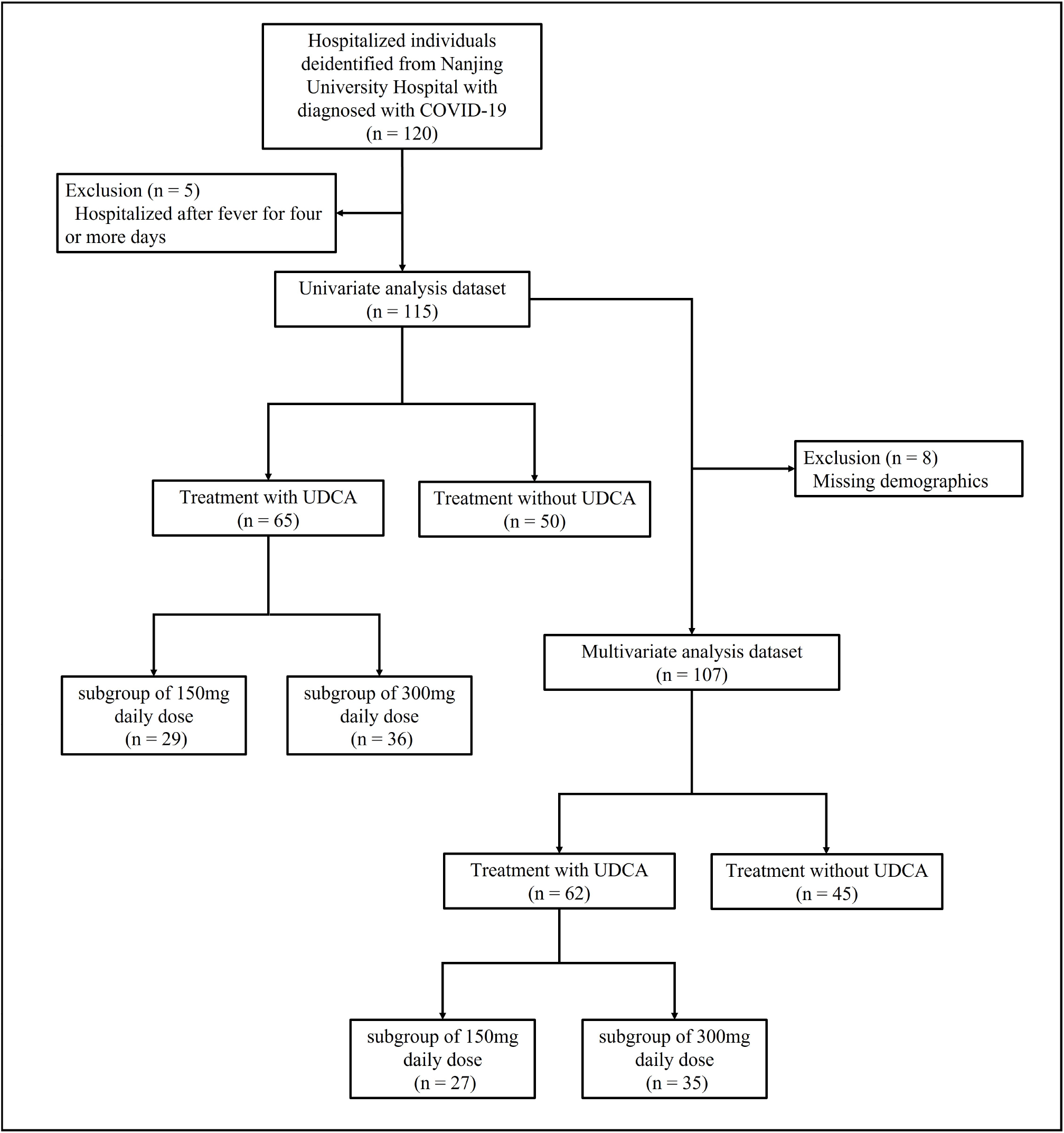
Study participant flowchart

**Extended Data Table 1.** Demographic and clinical characteristics by treatment group for UDCA

|  | with UDCA (N=65) | without UDCA (N=50) | P value |
| --- | --- | --- | --- |
| **Continuous variables** |  |  |  |
| Age, years |  |  | 0.698 |
| Median (IQR) | 23 (3.0) | 23 (3.8) |  |
| Missing data, no. (%) | 3 (4.6) | 4 (8.0) |  |
| BMI, kg/m<sup>2</sup> |  |  | 0.093 |
| Median (IQR) | 22.3 (4.6) | 20.9 (3.8) |  |
| Missing data, no. (%) | 3 (4.6) | 5 (10.0) |  |
| Body temperature on admission, °C |  |  | 0.175 |
| Median (IQR) | 39.3 (1.0) | 39.4 (0.8) |  |
| **Categorical variables** |  |  |  |
| Sex |  |  | 0.939 |
| Male, no. (%) | 40 (61.5) | 32 (64.0) |  |
| Female, no. (%) | 25 (38.5) | 18 (36.0) |  |
| Whether to use NSAIDs |  |  | <0.001 |
| Yes, no. (%) | 63 (96.9) | 38 (76.0) |  |
| No, no. (%) | 2 (3.1) | 12 (24.0) |  |
| Whether to use Symptomatic drugs |  |  | 0.316 |
| Yes, no. (%) | 64 (98.5) | 47 (94.0) |  |
| No, no. (%) | 1 (1.5) | 3 (6.0) |  |
| Days from fever onset to hospitalization |  |  | 0.214 |
| 0, no. (%) | 6 (9.2) | 2 (4.0) |  |
| 1, no. (%) | 18 (27.7) | 23 (46.0) |  |
| 2, no. (%) | 34 (52.3) | 21 (42.0) |  |
| 3, no. (%) | 7 (10.8) | 4 (8.0) |  |
| **Subgroup divided by a daily dose** |  |  | - |
| 150mg, no. (%) | 29 (44.6) | - |  |
| 300mg, no. (%) | 36 (55.4) | - |  |
| (All received at least two doses of the vaccine) |  |  |  |
| (All were youthful individuals with no comorbidities) |  |  |  |

**Extended Data Table 2.** Demographic and clinical characteristics by UDCA subgroup divided by daily dose

|  | **Daily 150mg**<br>**N=29** | **Daily 300mg or more**<br>**N=36** | ***P* value** |
| --- | --- | --- | --- |
| Continuous variables |  |  |  |
| Age, years |  |  | 0.109 |
| Median (IQR) | 24 (3.0) | 23 (3.0) |  |
| Missing data, no. (%) | 2 (6.8) | 1 (2.8) |  |
| BMI, kg/m<sup>2</sup> |  |  | 0.210 |
| Median (IQR) | 22.9 (3.2) | 21.3 (4.4) |  |
| Missing data, no. (%) | 2 (6.8) | 1 (2.8) |  |
| Body temperature on admission, °C |  |  | <0.001 |
| Median (IQR) | 38.8 (0.7) | 39.5 (0.5) |  |
| Categorical variables |  |  |  |
| Sex |  |  | 0.737 |
| Male, no. (%) | 19 (65.5) | 21 (58.3) |  |
| Female, no. (%) | 10 (34.5) | 15 (41.7) |  |
| Whether to use NSAIDs |  |  | 0.498 |
| Yes, no. (%) | 29 (100.0) | 34 (94.4) |  |
| No, no. (%) | 0 (0.0) | 2 (5.6) |  |
| Whether to use Symptomatic drugs |  |  | 1.000 |
| Yes, no. (%) | 29 (100.0) | 35 (97.2) |  |
| No, no. (%) | 0 (0.0) | 1 (2.8) |  |
| Days from fever onset to hospitalization |  |  | 0.650 |
| 0, no. (%) | 2 (6.9) | 4 (11.1) |  |
| 1, no. (%) | 10 (34.5) | 18 (22.2) |  |
| 2, no. (%) | 15 (51.7) | 19 (52.8) |  |
| 3, no. (%) | 2 (6.9) | 5 (13.9) |  |
| (All received at least two doses of the vaccine) |  |  |  |
| (All were youthful individuals with no comorbidities) |  |  |  |

**Extended Data Table 3.** Multivarite analysis of UDCA and no-UDCA groups after imputation

|  | With UDCA | Without UDCA |
| --- | --- | --- |
| No. of patients | 65 | 50 |
| No. of events (%) | 61 (93.8) | 38 (76.0) |
| Major adjusted with all covariates (Primary analysis) |  |  |
| Hazard ratio (95% CI) | 1.63 (1.01-2.60) | 1 (Reference) |
| P value | 0.044 |  |
| Unadjusted |  |  |
| Hazard ratio (95% CI) | 1.87 (1.06-3.32) | 1 (Reference) |
| P value | 0.031 |  |
| Minor adjusted with demographics, whether to use NSAIDs and days from fever onset to hospitalization |  |  |
| Hazard ratio (95% CI) | 1.66 (1.04-2.70) | 1 (Reference) |
| P value | 0.035 |  |
| Minor adjusted with demographics, whether to use NSAIDs and the temperature on admission |  |  |
| Hazard ratio (95% CI) | 1.64 (1.03-2.60) | 1 (Reference) |
| P value | 0.038 |  |

**Extended Data Table 4.** Multivarite analysis of subgroups after imputation

|  | Daily 150mg | Daily 300mg | Without UDCA |
| --- | --- | --- | --- |
| No. of patients | 29 | 36 | 50 |
| No. of events (%) | 28 (96.6) | 33 (91.7) | 38 (76.0) |
| Major adjusted with all covariates (Primary analysis) |  |  |  |
| Hazard ratio (95% CI) | 1.36 (0.76-2.40) | 1.86 (1.10-3.10) | 1 (Reference) |
| P value (Adjusted by B&H) | 0.293 (0.293) | 0.020 (0.040) |  |
| Unadjusted |  |  |  |
| Hazard ratio (95% CI) | 1.70 (0.86-3.50) | 2.00 (1.03-3.90) | 1 (Reference) |
| P value (Adjusted by B&H) | 0.125 (0.125) | 0.041 (0.081) |  |
| Minor adjusted with demographics, whether to use NSAIDs and days from fever onset to hospitalization |  |  |  |
| Hazard ratio (95% CI) | 1.49 (0.85-2.60) | 1.81 (1.08-3.10) | 1 (Reference) |
| P value (Adjusted by B&H) | 0.161 (0.161) | 0.025 (0.050) |  |
| Minor adjusted with demographics, whether to use NSAIDs and the temperature on admission |  |  |  |
| Hazard ratio (95% CI) | 1.39 (0.78-2.50) | 1.84 (1.11-3.10) | 1 (Reference) |
| P value (Adjusted by B&H) | 0.266 (0.266) | 0.019 (0.038) |  |

## Footnotes

1 Supplementary information will be available after peer-review.

2 Supplementary information will be available after peer-review.

## Notes

### Competing Interest Statement

The authors have declared no competing interest.

### Funding Statement

This study was funded by National Key Research and Development Program of China (2020AAA0107200), National Science Foundation of China (61921006), Jiangsu Provincial Natural Science Fund for Distinguished Young Scholars (BK20200005)

### Author Declarations

The Nanjing University Hospital institutional review boards approved this study as minimal risk and waived informed consent requirements.

